# Plaque characteristics derived from intravascular optical coherence tomography that predict major adverse cardiovascular events

**DOI:** 10.1101/2023.06.20.23291684

**Authors:** Juhwan Lee, Yazan Gharaibeh, Vladislav N. Zimin, Justin N. Kim, Issam Motairek, Neda S. Hassani, Luis A. P. Dallan, Gabriel T. R. Pereira, Mohamed H. E. Makhlouf, Ammar Hoori, Sadeer Al-Kindi, David L. Wilson

## Abstract

**Background:** With its near histological resolution and its optical contrast, intravascular optical coherence tomography (IVOCT) is the only imaging modality that allows a unique assessment of microscopic plaque characteristics. This study aimed to investigate whether plaque characteristics derived from IVOCT could predict a long-term major adverse cardiovascular event (MACE).

**Methods:** This study was a single-center, retrospective study on 104 patients who had undergone IVOCT-guided percutaneous coronary intervention. Plaque characterization was performed using OCTOPUS software developed by our group. A total of 31 plaque features, including lesion length, lumen, calcium, fibrous cap (FC), and vulnerable plaque features (e.g., microchannel and cholesterol crystal), were computed from the baseline IVOCT images (obtained before stenting). For IVOCT plaque features, the discriminatory power for predicting MACE was determined using univariate/multivariate logistic regression as assessed by area under the receiver operating characteristic curve (AUC).

**Results:** Of 104 patients, MACE was identified in 24 patients (23.1%). Univariate logistic regression revealed that lesion length, maximum calcium angle, maximum calcium thickness, maximum FC angle, maximum FC area, and FC surface area were significantly associated with MACE (p<0.05). Additionally, cholesterol crystal and layered plaque showed a strong association with MACE (p<0.05). In the multivariate logistic analysis, only the FC surface area (OR 2.38, CI 0.98-5.83, p<0.05) was identified as a significant determinant for MACE, highlighting the importance of the 3D lesion analysis. The AUC of FC surface area for predicting MACE was 0.851 (95% CI 0.800-0.927, p<0.05). Luminal stenosis was not a strong predictor of the risk of MACE.

**Conclusions:** Patients with MACE had distinct plaque characteristics in IVOCT. In particular, large FC surface areas were a risk factor. Interestingly, cap thickness, a commonly highlighted feature for lesion vulnerability, was less predictive than cap area. Studies such as this one might someday lead to recommendations for pharmaceutical and interventional approaches.

## Introduction

Coronary artery disease (CAD) continues to be a significant cause of morbidity and mortality on a global scale ^1^. Given the prevalence of atherosclerosis in a substantial proportion of the population ^2^, there is a growing interest in accurately assessing disease aggressiveness by predicting major adverse cardiovascular events (MACE) through medical imaging. Various imaging modalities have been employed to visualize plaque in coronary vessels and characterize CAD. In coronary computed tomography angiography (CCTA) images, several high-risk features have been identified, such as positive remodeling, low attenuation plaque, napkin-ring sign, and spotty calcification, with their association to MACE serving as a key determinant ^3–6^. Notably, recent research has emphasized the assessment of pericoronary fat in CCTA images, including radiomics evaluations, once again leveraging their association with MACE to support the rationale ^5, 7–12^. Using CT calcium score images, one can predict MACE using the Agatston score computed from calcifications ^13–15^.

None of the non-invasive imaging modalities currently available can match the exceptional resolution and contrast achieved with intravascular optical coherence tomography (IVOCT) images ^16^. Notably, IVOCT provides superior resolution (axial: 10 *μm*, lateral: 20-40 *μm*), allowing for precise identification of thin-cap fibroatheroma (TCFA). Through IVOCT imaging, we can quantitatively assess the microscopic characteristics of plaque, including TCFA, macrophage infiltration, cholesterol crystal presence, and microchannels, as validated by histological records ^17–36^. Furthermore, histopathological investigations have indicated a strong association between the pathogenesis of most acute coronary events (such as plaque rupture and myocardial infarction) and the presence of microcalcification, TCFA, and large lipid-rich necrotic cores ^37–39^. These plaques are also often characterized by both intraplaque and inflammation that are strongly associated with plaque progression ^40^. Extensive ex vivo and in vitro studies have further substantiated the correlation between macrophage infiltration and vulnerable plaque characteristics through histopathological evidence ^37, 38, 41–43^. As IVOCT gives a promising, unique perspective on coronary plaque, there is a strong rationale to relate findings from IVOCT on a long-term cardiovascular risk assessed by the probability for MACE.

To date, no studies have investigated whether plaque characteristics derived from IVOCT imaging can serve as predictors of MACE. We hypothesize that plaque characteristics derived from IVOCT are related to the presence of MACE. Specifically, we aim to identify relevant features in pre-stent IVOCT images that exhibit an association with MACE. This analysis will not only unveil high-risk features extracted from IVOCT but will also enable subsequent correlative investigations with CCTA to identify novel high-risk CCTA features based on their association with IVOCT findings. In this regard, we have found a correlation between specific radiomic features of pericoronary fat in CCTA images and microscopic IVOCT characteristics, including thin-cap fibroatheroma (TCFA) and microchannels ^44^. It is important to note that high-risk features identified through their association with MACE may differ from those associated with risk in a specific lesion, as identified solely by CCTA.

In this study, we investigated the association between plaque characteristics observed in IVOCT imaging and the occurrence of MACE. Using OCTOPUS software ^45^ on IVOCT images, we segmented plaques into constituent parts (e.g., calcium and fibrous cap (FC)), extracted IVOCT plaque features (e.g., calcium thickness and FC surface area), and determined their discriminatory power for predicting MACE. Notably, this analysis specifically focused on microscopic features that are exclusively visible through IVOCT, such as FC and microchannel components.

## Methods

### Study population

We retrospectively reviewed 805 patients and enrolled 104 patients with coronary artery disease who had undergone clinically indicated invasive coronary X-ray angiography and IVOCT-guided percutaneous coronary intervention (PCI) at the University Hospitals Cleveland Medical Center in Cleveland, Ohio, USA, between 2013 and 2019. We included patients who had a culprit lesion identified on coronary X-ray angiography. Exclusion criteria consisted of ostial lesion, inability to cross the lesions with the OCT catheter due to the tortuosity and/or occluding thrombus, bypass graft stenosis, in-stent restenosis, and chronic total occlusion. This study was conducted in compliance with the Declaration of Helsinki and received approval from the Institutional Review Board of University Hospitals Cleveland Medical Center (Cleveland, Ohio, USA).

### IVOCT imaging and plaque characterization

Invasive coronary angiography was conducted using 6-7 Fr catheters via radial or femoral access, following the administration of 250 *µg* of intracoronary nitroglycerine. The obtained coronary angiogram was analyzed using QAngio® software (Medis, Leiden, the Netherlands). IVOCT-guided PCI was performed employing conventional techniques. During the procedure, the interventional cardiologist exercised discretion in selecting stenting variables such as stent length and diameter. Only drug-eluting stents were utilized in this study. IVOCT images were acquired using the C7XR FD-OCT Imaging system (Abbott Vascular, Santa Clara, CA, USA), following the administration of nitroglycerin (100-200*g*). To reach the lesion of interest, a 2.7-Fr OCT catheter (Dragonfly OPTIS, Abbott Vascular, Santa Clara, CA, USA) was advanced over a conventional guidewire, with the catheter position verified through invasive coronary angiography. Non-diluted iodine contrast (ISOVUE-370, iopamidol injection, 370 *mg* iodine/mL; Bracco Diagnostics Inc., Princeton, NJ, USA) was employed to achieve blood clearance. Subsequently, imaging pullback was executed at a frame rate of 180 fps, a pullback speed of 36 *mm/s*, and an axial resolution of approximately 20 *µm*. The acquired images were deidentified and forwarded to the Core Laboratory for independent offline analysis.

Plaque and vessel analysis was conducted using the Optical Coherence TOmography PlaqUe and Stent (OCTOPUS) software, which was previously developed and validated by our research group ^45^. In this study, the OCTOPUS software automatically segmented the lumen, calcified plaque, lipidic plaque, and FC. Additionally, the presence of vulnerable plaques, including microchannels, macrophage infiltration, cholesterol crystals, layered plaques, and calcium nodules, was assessed using the OCTOPUS software. If necessary, manual editing of the results was performed according to the definitions provided in the “consensus document” ^46^.

### IVOCT feature extraction

We analyzed 31 IVOCT features from the baseline IVOCT images taken prior to stenting to predict the occurrence of MACE. The features were automatically computed using OCTOPUS software ^45^, except for the vulnerable plaque features. Lesion length was defined as the length of the vessel segment where the stent was deployed. Lumen features included minimum/average lumen area and minimum/average lumen diameter. Calcium features encompassed maximum/minimum calcium angle, thickness, and depth. FC features were evaluated using four categories of FC thickness (1: thickness ≤65 *µm*, 2: 65 *µm* < thickness < 150 *µm*, 3: thickness ≥150 *µm*, and T: total) and included maximum/minimum FC angle, thickness, area, surface area, and burden. For example, FC surface area-1 represented the surface area of FC regions with a thickness ≤ 65 µm, while FC surface area-T represented the total surface area of FC regions, regardless of thickness. FC angle, thickness, and area were calculated from each IVOCT image frame, while FC surface area and burden were computed for the entire lesion. FC surface area was defined as the total area covered by the FC on the surface of the vessel lumen, as visualized in the en face view (*θ,z*), while FC burden was calculated as the ratio of FC area to the surface area of the vessel lumen. Vulnerable plaque features included the presence of microchannels, macrophage infiltration, cholesterol crystals, layered plaque, or calcium nodules in the lesion. Table 1 provides a summary of the IVOCT features used in this study for predicting MACE.

**Table 1.**
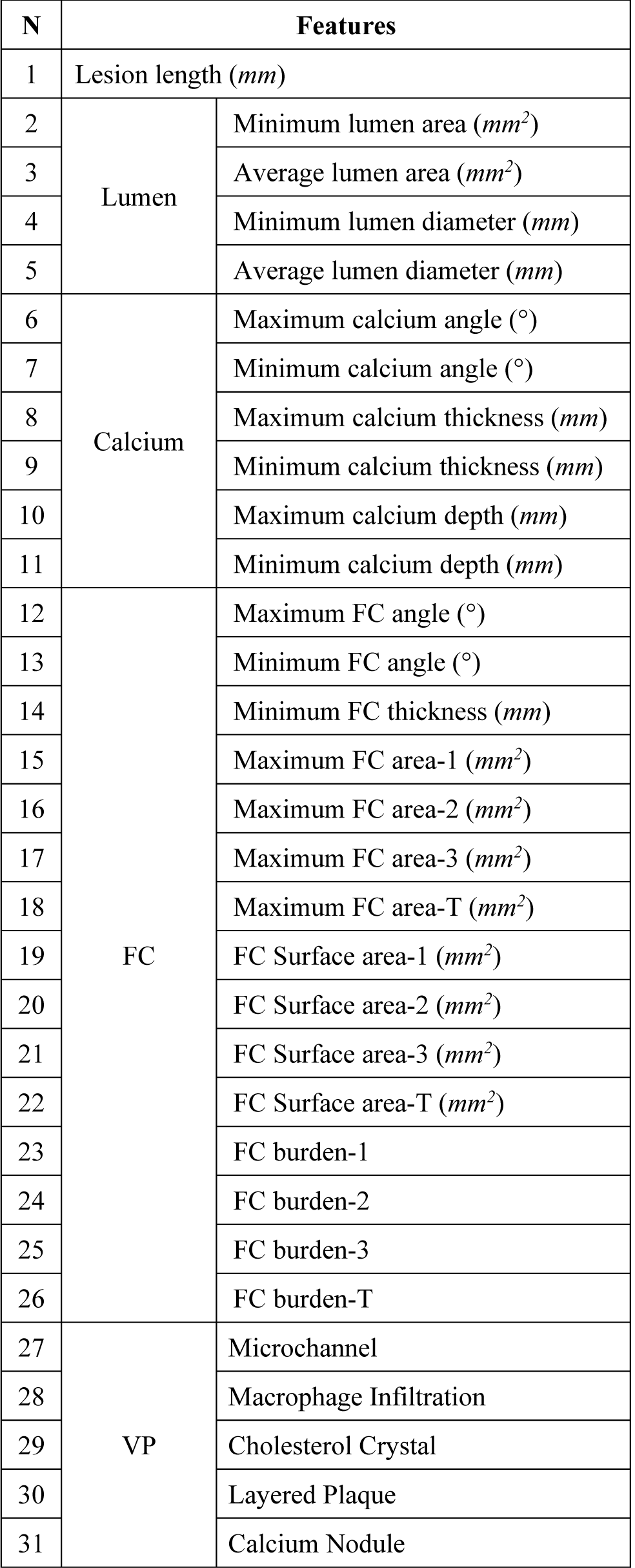
IVOCT plaque features, including lesion length, and feature groupings (4 lumen, 6 calcium, 15 FC, and 5 vulnerable plaque features). FC refers to the fibrous cap, and VP represents vulnerable plaque.

### Clinical endpoint

Details regarding the occurrence of MACE were collected from the electronic medical records of the University Hospitals Cleveland Medical Center, located in Cleveland, OH, USA. MACE was defined as cardiovascular death resulting from acute myocardial infarction, heart failure, stroke, and other cardiovascular causes.

### Statistical analysis

We conducted various analyses on the IVOCT plaque features. Continuous features were presented as mean ± standard deviation, while categorical features were reported as frequencies. Statistical comparisons between the MACE and no-MACE groups were performed using a student t-test. To assess the inter-correlations of IVOCT plaque features, a heatmap analysis was conducted using the non-parametric Spearman’s rank correlation coefficient and hierarchical clustering. For the prediction of MACE, both univariate and multivariate logistic regression models were employed, with 95% confidence intervals (CI) calculated. In the multivariate logistic regression, features that showed significance (p<0.05) in the univariate analysis were included. The discriminatory power of the models for predicting MACE was evaluated using the area under the receiver operating characteristic curve (AUC). The optimal cutoff values on the ROC curve were determined based on the maximum sum of sensitivity and specificity. Statistical significance was defined as a p-value less than 0.05. All statistical analyses were performed using R Studio software (version 1.4.1717, R Foundation for Statistical Computing, Vienna, Austria).

## Results

This study comprised 104 patients with coronary artery disease who underwent IVOCT-guided PCI. No patients were excluded based on clinical characteristics. Among the 104 patients, the mean age was 67.1±12.0 years, with 74 males (71.2%). During an average follow-up period of 19 months, MACE occurred in 24 patients. Among the study population, 99 patients (95.2%) had hypertension, 53 patients (51.0%) were current smokers, and 56 patients (53.8%) had diabetes mellitus. The baseline characteristics of the study population are presented in Table 2.

**Table 2.**
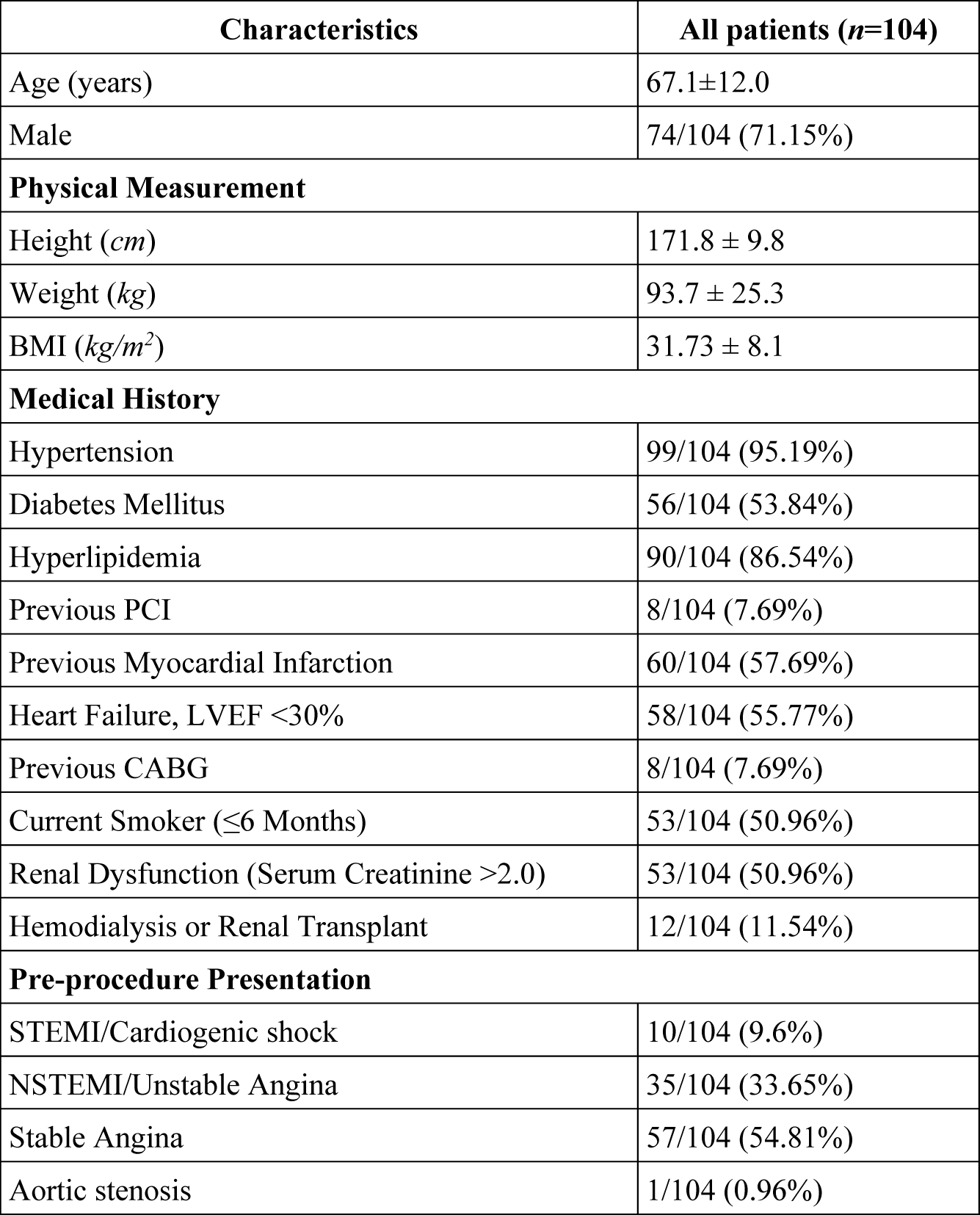
Baseline characteristics of the study population.

We conducted a comparative analysis of plaque features between the MACE and no-MACE groups. The MACE group exhibited significantly larger lesion length, maximum calcium angle, and maximum calcium thickness compared to the no-MACE group (p<0.05 for all) (Table 3). Similarly, there was a significant association between the occurrence of MACE and increasing FC features, including maximum FC angle, maximum FC area, FC surface area, and FC burden, as compared to the no-MACE group. Notably, maximum FC area-T and FC surface area-T demonstrated the smallest p-values (p<0.000001). However, quantitative features related to lumen, calcium depth, and minimum FC thickness did not show significant differences between the MACE and no-MACE groups. Furthermore, all vulnerable plaque features, such as microchannel and cholesterol crystal, were more frequently observed in the MACE group compared to the no-MACE group. Table 3 provides a comprehensive comparison of IVOCT plaque features using a student t-test.

**Table 3.** Comparison of IVOCT plaque features between MACE and no-MACE groups using a t-test.

| Features | MACE (n=24) | No-MACE (n=80) | P-value |
| --- | --- | --- | --- |
| Lesion length (mm) | 37.15±14.15 | 28.56±11.51 | <0.05 |
| Maximum calcium angle (°) | 245.19±83.08 | 162.60±71.08 | <0.001 |
| Minimum calcium angle (°) | 20.31±12.22 | 15.86±5.24 | 0.06 |
| Maximum calcium thickness (mm) | 1.52±0.24 | 1.24±0.29 | <0.001 |
| Minimum calcium thickness (mm) | 0.30±0.10 | 0.27±0.06 | 0.28 |
| Maximum calcium depth (mm) | 0.52±0.18 | 0.50±0.19 | 0.81 |
| Minimum calcium depth (mm) | 0.008±0.010 | 0.014±0.014 | 0.22 |
| Minimum lumen area (mm<sup>2</sup>) | 2.03±1.04 | 2.07±1.25 | 0.91 |
| Average lumen area (mm<sup>2</sup>) | 5.60±2.37 | 5.87±2.60 | 0.72 |
| Minimum lumen diameter (mm) | 0.96±0.28 | 1.00±0.37 | 0.75 |
| Average lumen diameter (mm) | 2.57±0.56 | 2.62±0.56 | 0.75 |
| Maximum FC angle (°) | 142.00±51.21 | 61.40±53.63 | <0.00001 |
| Minimum FC angle (°) | 29.81±11.75 | 28.29±27.06 | 0.83 |
| Minimum FC thickness (mm) | 0.0228±0.0095 | 0.0409±0.0567 | 0.21 |
| Maximum FC area-1 (mm<sup>2</sup>) | 0.48±0.29 | 0.14±0.21 | <0.00001 |
| Maximum FC area-2 (mm<sup>2</sup>) | 1.73±0.75 | 0.62±0.67 | <0.000001 |
| Maximum FC area-3 (mm<sup>2</sup>) | 1.38±0.64 | 0.60±0.65 | <0.0001 |
| Maximum FC area-T (mm<sup>2</sup>) | 3.60±1.30 | 1.35±1.23 | <0.0000001 |
| FC Surface area-1 (mm<sup>2</sup>) | 0.51±0.49 | 0.07±0.13 | <0.00001 |
| FC Surface area-2 (mm<sup>2</sup>) | 5.36±5.53 | 0.72±1.08 | <0.00001 |
| FC Surface area-3 (mm<sup>2</sup>) | 3.77±3.76 | 0.58±1.09 | <0.00001 |
| FC Surface area-T (mm<sup>2</sup>) | 9.63±8.73 | 1.37±1.81 | <0.000001 |
| FC burden-1 | 42.30±42.10 | 5.56±9.81 | <0.00001 |
| FC burden-2 | 496.35±796.34 | 53.59±78.55 | <0.001 |
| FC burden-3 | 369.09±570.24 | 40.31±63.43 | <0.001 |
| FC burden-T | 907.75±1368.83 | 99.46±121.49 | <0.001 |
| Microchannel | 9 (37.5%) | 11 (16.7%) |  |
| Macrophage Infiltration | 21 (87.5%) | 50 (78.6%) |  |
| Cholesterol Crystal | 17 (68.8%) | 12 (19.1%) |  |
| Layered Plaque | 8 (31.3%) | 3 (4.8%) |  |
| Calcium Nodule | 8 (31.3%) | 8 (11.9%) |  |

In order to address the issue of correlated features, we conducted a hierarchical clustering analysis of IVOCT plaque features using the non-parametric Spearman’s rank correlation coefficient (Fig. 1). Among all the IVOCT plaque features, we identified 12 features (minimum lumen diameter, average lumen diameter, maximum FC area 1/2/3, FC surface area 1/2/3, and FC burden 1/2/3/T) with Spearman’s correlation coefficients exceeding 0.9. Consequently, we reduced the total number of features extracted from the IVOCT images from 31 to 14, considering only features with a rho-value greater than 0.9. Notably, within the heatmap, the FC features exhibited the smallest p-values, indicating their potential significance in relation to MACE prediction.

**Figure 1.**
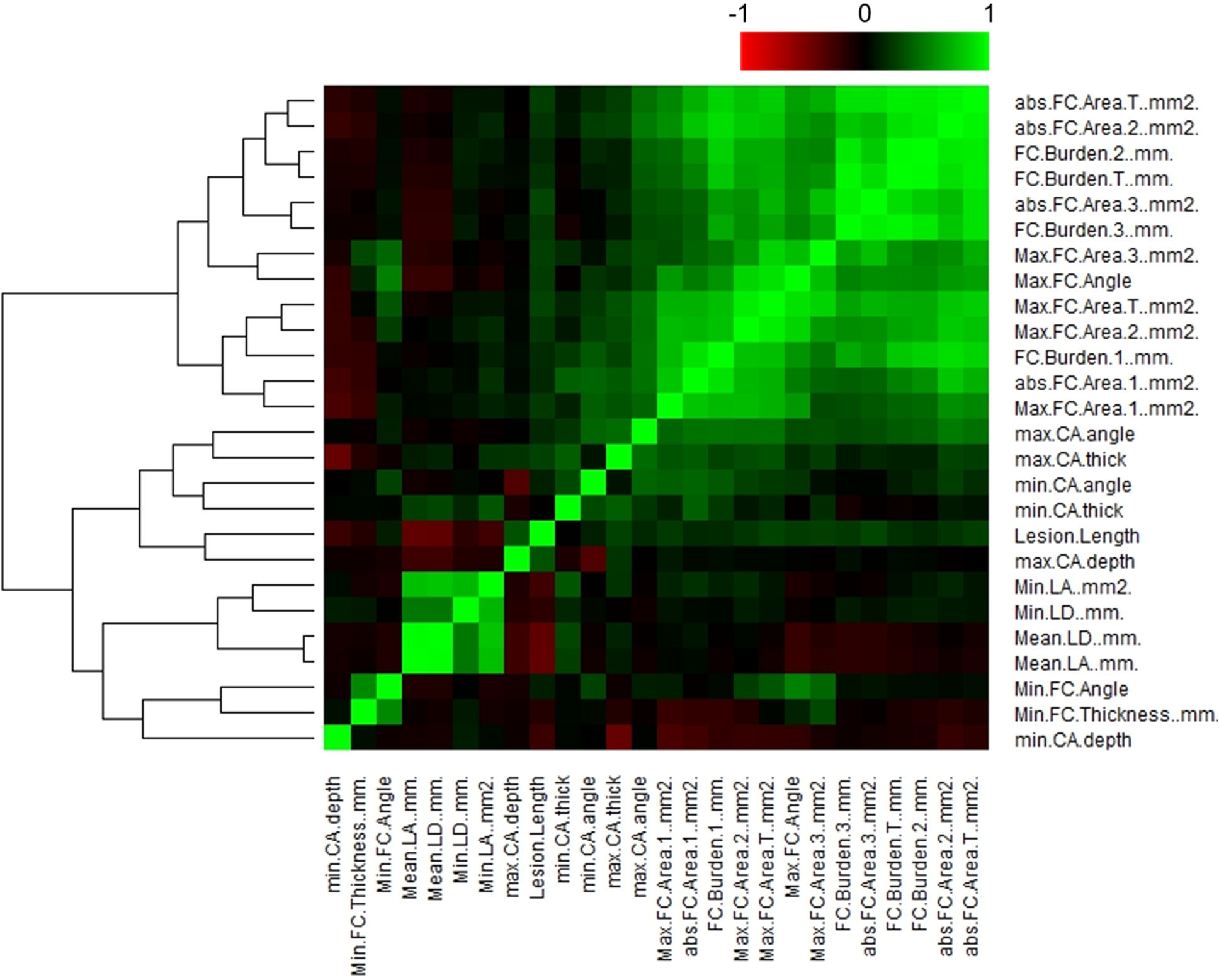
Correlation plot of IVOCT plaque features, illustrating hierarchical clustering and distinct clusters of feature correlation. The correlation coefficient (*R*) values are plotted against each other. The color key represents the explained variances: *R* values < 0.5 are shown in black, while greater values are depicted in green or red with increasing intensity. Using a Spearman correlation coefficient threshold of 0.9, a total of 12 features, including minimum/mean lumen diameter, maximum FC area-1/2/3, FC surface area-1/2/3, and FC burden-1/2/3/T, were excluded from further analysis.

To assess the discriminatory ability of the features in predicting MACE events, we performed univariate and multivariate logistic regression analyses. In the univariate regression analysis, several features including lesion length, maximum calcium angle, maximum calcium thickness, maximum FC angle, maximum FC area-T, FC surface area-T, cholesterol crystal, and layered plaque demonstrated significant associations with MACE (Table 4). On the other hand, minimum/average lumen area and FC thickness did not independently predict MACE. Subsequently, in the multivariate analysis, only the FC surface area-T (OR 2.38, CI 0.98-5.83, p<0.05) exhibited a strong association with the occurrence of MACE (Table 4). The IVOCT plaque features that showed significant associations with MACE in both univariate and multivariate logistic regression analyses are summarized in Table 4.

**Table 4.** Univariate/multivariate logistic regression for predicting MACE. Eight IVOCT plaque features showed significant correlation with MACE in the univariate logistic regression analysis. In the multivariate logistic regression analysis, only the FC surface area-T demonstrated a strong association with a higher prevalence of MACE (p<0.05).

| Features | Univariate Logistic Regression |  |  |  | Multivariate Logistic Regression |  |  |  |
| --- | --- | --- | --- | --- | --- | --- | --- | --- |
|  | P-value | Odd Ratio | Lower 95% | Upper 95% | P-value | Odd Ratio | Lower 95% | Upper 95% |
| Lesion length (*mm*) | <0.05 | 1.05 | 1.01 | 1.11 | 0.90 | 0.99 | 0.89 | 1.11 |
| Maximum calcium angle (°) | <0.05 | 1.00 | 1.01 | 1.01 | 0.29 | 1.01 | 0.99 | 1.02 |
| Minimum calcium angle (°) | 0.09 | 1.07 | 0.99 | 1.16 |  |  |  |  |
| Maximum calcium thickness (*mm*) | <0.05 | 48.48 | 3.82 | 614.87 | 0.06 | 190.55 | 0.73 | 4993.57 |
| Minimum calcium thickness (*mm*) | 0.29 | 62.40 | 0.03 | 1224.3 |  |  |  |  |
| Maximum calcium depth (*mm*) | 0.81 | 1.46 | 0.07 | 31.17 |  |  |  |  |
| Minimum calcium depth (*mm*) | 0.07 | 0.02 | 0.00 | 1.30 |  |  |  |  |
| Minimum lumen area (*mm*<sup>2</sup>) | 0.90 | 0.97 | 0.59 | 1.59 |  |  |  |  |
| Average lumen area (*mm*<sup>2</sup>) | 0.71 | 0.96 | 0.75 | 1.21 |  |  |  |  |
| Maximum FC Angle (°) | <0.05 | 1.03 | 1.01 | 1.05 | 0.10 | 1.05 | 0.99 | 1.12 |
| Minimum FC Angle (°) | 0.83 | 1.00 | 0.98 | 1.03 |  |  |  |  |
| Minimum FC Thickness (*mm*) | 0.23 | 0.00 | 0.00 | 920.14 |  |  |  |  |
| Maximum FC Area-T (*mm*<sup>2</sup>) | <0.001 | 5.65 | 2.03 | 15.69 | 0.16 | 0.14 | 0.01 | 2.13 |
| FC Surface Area-T (*mm*<sup>2</sup>) | <0.001 | 2.08 | 1.42 | 3.04 | <0.05 | 2.38 | 0.98 | 5.83 |
| Microchannel | 0.10 | 3.00 | 0.82 | 10.98 |  |  |  |  |
| Macrophage Infiltration | 0.44 | 1.91 | 0.36 | 9.99 |  |  |  |  |
| Cholesterol Crystal | <0.001 | 9.35 | 2.53 | 34.58 | 0.42 | 3.22 | 0.19 | 54.85 |
| Layered Plaque | <0.05 | 9.09 | 1.55 | 53.39 | 0.48 | 5.18 | 0.06 | 489.67 |
| Calcium Nodule | 0.09 | 3.36 | 0.82 | 13.78 |  |  |  |  |

Using the single best feature, which was determined to be FC surface area-T based on the multivariate analysis, we constructed an ROC curve to assess the predictive capability of the IVOCT plaque feature for MACE (Fig. 2). FC surface area-1 made the smallest contribution, while FC surface area-2 (65 *µm*<T<150 *µm*) contributed the most to MACE prediction. The combined AUC for FC surface area-T was 0.851 (95% CI 0.800-0.927). A significant difference in FC surface area-T was observed using a box plot analysis (Fig. 3). Additionally, Figure 4 presents a 3D visualization of high-risk and low-risk lesions in representative IVOCT pullbacks. The high-risk lesion exhibited a thicker calcification thickness (1.32 *mm*) and a larger FC surface area (39.10 *mm*^2^), whereas the low-risk lesion had a thinner calcium thickness (0.72 *mm*) and a smaller FC surface area (1.63 *mm*^2^).

**Figure 2.**
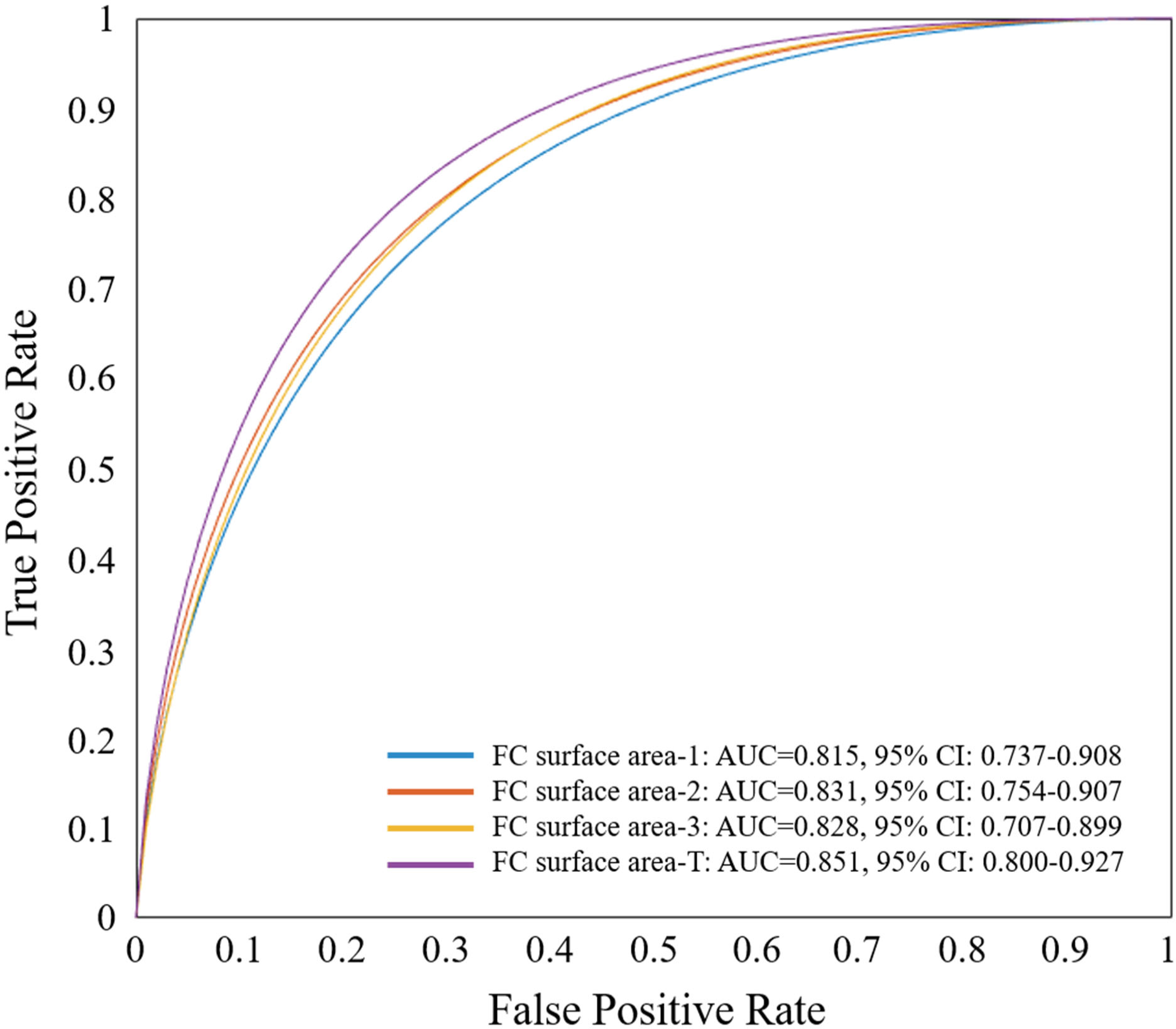
ROC curve analysis of the most significant IVOCT plaque feature, FC surface area-T, determined by multivariate logistic regression for predicting MACE. ROC curves of FC surface area 1-3 are also displayed. The FC area with T<65 *µm* contributed the least (AUC: 0.815, 95%CI: 0.737-0.908), while the FC area with 65 *µm*<T<150 *µm* contributed the most (AUC: 0.831, 95% CI: 0.754-0.907) in predicting MACE. When combined, FC surface area-T had an AUC of 0.851 (95% CI 0.800-0.927).

**Figure 3.**
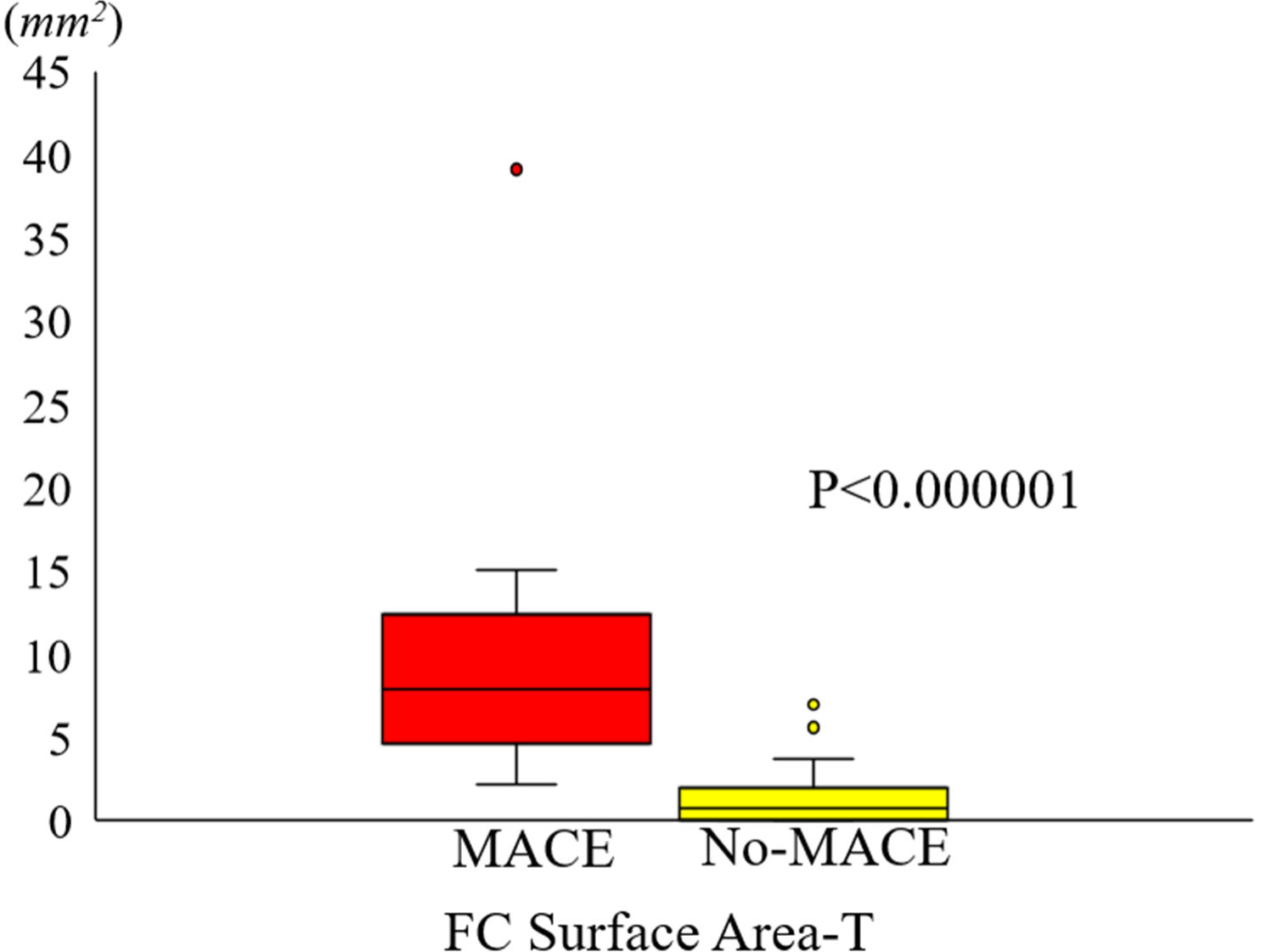
Box plot illustrating the disparity in FC surface area-T between the MACE and no-MACE groups. The FC surface area-T in the MACE group (9.63±8.73 *mm*^2^) was significantly greater than that in the no-MACE group (1.37±1.81 *mm*^2^) (p<0.000001). The MACE group is represented in red, while the no-MACE group is depicted in yellow.

**Figure 4.**
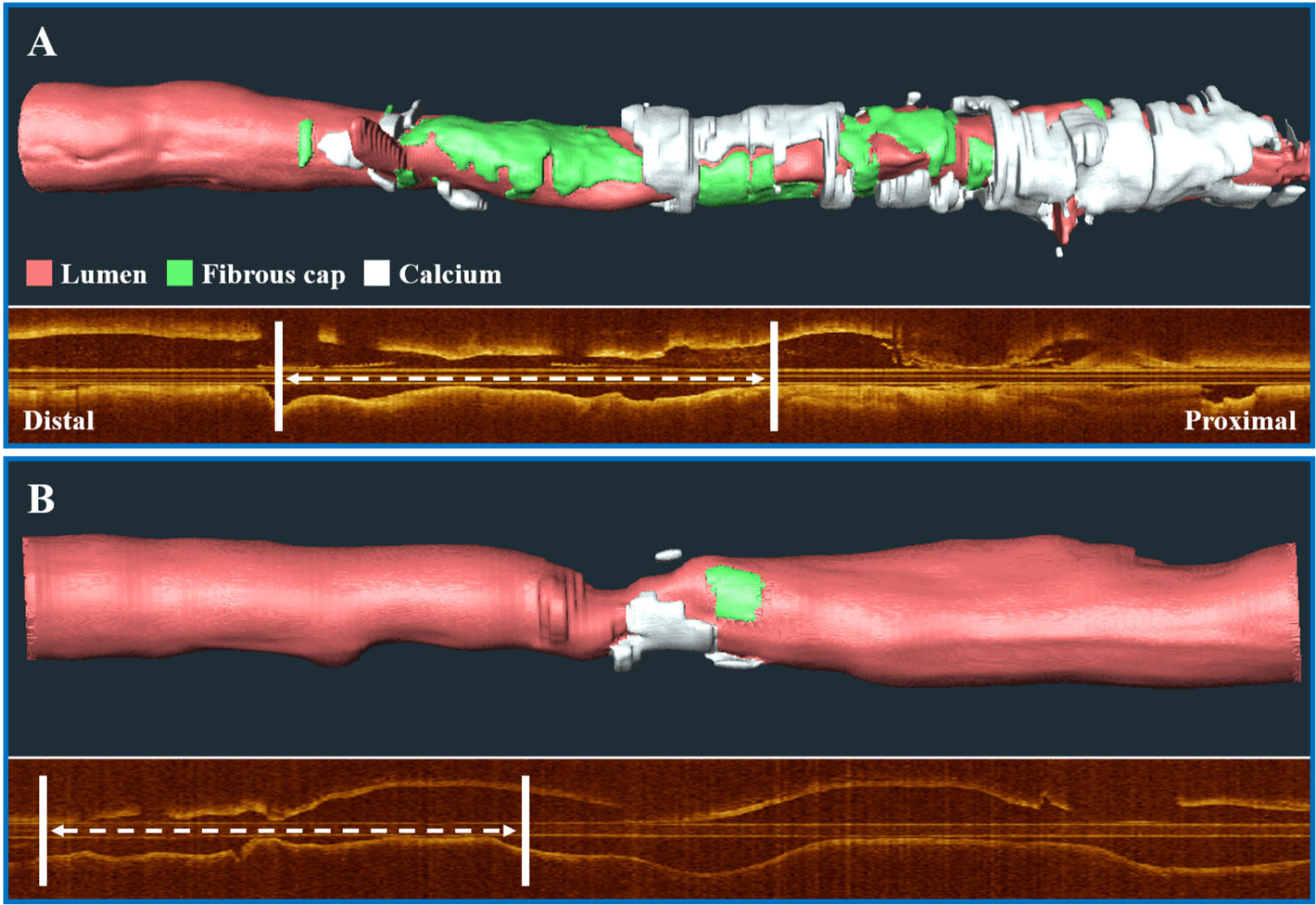
Three-dimensional (3D) visualization of high-risk and low-risk lesions on representative IVOCT pullbacks. The top panel (A) shows a high-risk lesion with a maximum calcium thickness of 1.32 *mm* and an FC surface area of 39.10 *mm*^2^. The bottom panel (B) displays a low-risk lesion with a maximum calcium thickness of 0.72 *mm* and an FC surface area of 1.63 *mm*^2^. Subsequently, the high-risk case experienced a MACE, while the low-risk case remained event-free. The corresponding longitudinal IVOCT maps are provided below each case, with white arrows indicating the location of the lesion where the stent was implanted. The lumen is represented in red, the FC in green, and the calcium in white.

## Discussion

To the best of our knowledge, this study represents the first investigation into the predictive potential of IVOCT-derived plaque characteristics for future MACE. Building upon our previous research utilizing IVOCT imaging ^45, 47–60^, we aimed to establish correlations between IVOCT plaque features and the occurrence of MACE. This study offers several noteworthy contributions. First, we employed our interactive OCTOPUS software to automatically compute plaque features such as FC thickness and FC surface area. Second, our analysis of features associated with MACE revealed the presence of high-risk features, including FC surface area, as well as unexpectedly low-risk features, such as TCFA thickness, when assessed in the context of stent-treated lesions. Third, preliminary ROC analysis indicates that features observed in IVOCT images hold the potential to predict future adverse events, potentially offering insights into patient management strategies.

The predictive power of FC surface area surpassed that of FC thickness, which has been a commonly studied risk factor for lesion rupture in previous reports ^41, 61, 62^. Our findings highlight that lesion area plays a more critical role in predicting MACE. This may be partially attributed to temporal trends in plaque pathogenesis. Typically, the formation of a plaque is preceded by the accumulation of lipid-laden macrophages, which later undergo apoptosis, resulting in the development of a necrotic lipid-rich core. Subsequently, fibrous tissue forms in the intima, leading to the formation of a fibrous cap. Thinning of the fibrous cap typically occurs after plaque enlargement ^63^. Therefore, the “3D” lesion size may serve as an earlier indicator of plaque instability compared to the “later” appearance of a localized thin fibrous cap. Considering the diffuse inflammatory nature of atherosclerosis, it is crucial to analyze the entire 3D lesion, specifically focusing on FC surface area. In another study, we also observed that FC surface area was a strong indicator for predicting the development of neo-atherosclerosis ^64^. In contrast, no assessment of thin cap was a strong predictor of MACE. That is, neither minimum thickness nor FC surface area-1 were highly predictive. Nevertheless, it is important to note that the significance of FC thickness should not be overlooked. In our analysis, FC thickness was obtained from a lesion that was subsequently treated with a stent, providing additional protection to the lesion. MACE is unlikely to be associated with the treated lesion, although in-stent thrombosis and restenosis are possible. It is likely that our assessments are associated with the extent of disease. While a thin fibrous cap may signify local plaque instability, a larger surface area may indicate a more widespread burden of atherosclerotic disease. This could potentially translate into acute events occurring in regions outside the imaged plaque, maybe even in another vessel.

Interestingly, luminal stenosis (minimum lumen area) was not a strong predictor of the risk of MACE. That is, the severity of a treated, flow limiting stenosis is not a reliable indicator of future MACE. Based on the arguments presented above, it does not seem to be a predictor of MACE-related disease in the rest of the heart. This is to be expected, as a more comprehensive 3D analysis of the vessel provides a more reliable assessment for MACE, taking into account numerous variable interactions, unlike a single measurement like the minimum lumen area. This finding aligns with a study conducted by Kim et al. ^65^, which found no significant differences in minimal lumen diameter and percentage of stenosis between lesions with and without adverse clinical outcomes in patients who underwent IVOCT-guided PCI, thus further supporting our results.

Long-term outcome prediction studies have predominantly utilized CCTA and cardiac magnetic resonance angiography (CMRA). These studies have focused on clinical characteristics and high-risk plaque features such as spotty calcification, low-attenuation plaque, positive remodeling, and the napkin-ring sign ^66, 67^. Recently, there has been a growing number of studies that aim to predict long-term outcomes using radiomic features in CCTA ^5, 7–12^. For instance, Oikonomou et al. calculated a total of 843 radiomic features (including shape-related, first-order, and texture features) in CCTA images and correlated them with the occurrence of MACE within 5 years of the CCTA scan ^9^. They discovered a high-risk radiomic profile of pericoronary fat that may be associated with an elevated cardiac risk. Similarly, Kolossváry et al. analyzed 935 radiomic features, including first-order and textural features with different bin sizes, to identify invasive and radionuclide imaging markers of plaque vulnerability ^5^. Their results demonstrated that the most informative radiomic features were able to identify attenuated plaque as observed by intravascular ultrasound (IVUS), TCFA as observed by IVOCT, and NaF18-positivity. However, non-invasive imaging modalities such as CCTA and CMRA only have a moderate correlation with the gold standard intravascular imaging techniques (e.g., IVOCT) and provide limited information about the artery wall and microscopic features of atherosclerosis. IVOCT, with its nearly histological resolution (axial: 10 *μm*, lateral: 20-40 *μm*) and optical contrast, offers a comprehensive assessment of coronary arteries ^16^. Specifically, it enables unique evaluation of microscopic plaque components such as FC, macrophages, cholesterol crystals, and microchannels, in addition to macroscopic plaques including fibrous and calcified plaques. Although IVOCT provides a better representation of the state of atherosclerosis compared to non-invasive imaging modalities, no studies have quantitatively analyzed IVOCT plaque characteristics for predicting future adverse outcomes. In this study, we utilized OCTOPUS ^45^ to perform plaque characterization in IVOCT images, and for the first time, identified FC surface area as a significant determinant of long-term MACE outcomes. Our results are promising and have the potential to optimize treatment strategies and improve short- and long-term outcomes.

Improved and automated characterization of atherosclerosis in IVOCT holds the potential to enable personalized treatments. The advancements in preventive and cardioprotective therapeutics over the past decade, including P2Y12 antagonists, direct oral anticoagulants, proprotein convertase subtilisin/kexin 9 (PCSK9) inhibitors, icosapent ethyl, glucagon-like peptide 1 (GLP-1) agonists, and others, highlight the need for personalized medicine approaches that ensure appropriate treatment for individual patients in a cost-effective manner. Automated identification of high-risk vessels would provide an opportunity to guide the implementation of intensive therapies in clinical practice and enhance patient cohorts for testing the effectiveness of emerging novel therapeutics. Furthermore, accurate identification of high-risk lesions could inform potential revascularization strategies. For instance, in addition to treating a stenosis, an extra stent could be added to seal a high-risk lesion. The assessment of plaque changes with precise registration has the potential to facilitate mechanistic studies in drug development ^68^. Additionally, the identification of high-risk IVOCT characteristics could offer insights to other imaging modalities.

This study has some limitations that should be acknowledged. First, it was a retrospective study conducted at a single center, and the sample size was relatively small. This may limit the generalizability of the findings to a larger population. Second, the study focused specifically on patients undergoing PCI with available IVOCT data. It remains unclear whether the identified features would be applicable and relevant to a more diverse and broader population.

In conclusion, our study demonstrates that patients with MACE have distinct plaque characteristics in IVOCT images compared to those without MACE. Particularly, FC surface area showed a strong predictive value, while features related to cap thickness were less predictive, despite their emphasis in the literature regarding lesion vulnerability. These findings have potential implications for patient management, allowing identification of individuals at higher risk for future events. Furthermore, correlating the features identified in our study with those observed in other imaging modalities, such as CCTA, through multi-modality imaging studies could provide valuable insights.

## Data Availability

The datasets generated and/or analyzed during the current study are not publicly available due to legal/ethical reasons but are available from the corresponding author on reasonable request.

## Acknowledgments

The content of this report is solely the responsibility of the authors and does not necessarily represent the official views of the National Institutes of Health. The grants were obtained via collaboration between Case Western Reserve University and University Hospitals Cleveland Medical Center. This work made use of the High-Performance Computing Resource in the Core Facility for Advanced Research Computing at Case Western Reserve University. The veracity guarantor, Justin N. Kim, affirms to the best of his knowledge that all aspects of this paper are accurate.

